# Prevalence and Determinants of Prehospital Impression of Stroke in Ischemic Stroke Patients

**DOI:** 10.1101/2024.09.25.24314407

**Authors:** Mauro Caffarelli, Andrew J. Wood, Remle P Crowe, Edilberto Amorim, Hooman Kamel, Anthony S. Kim, Elan L. Guterman

## Abstract

**BACKGROUND:** Emergency Medical Services (EMS) clinicians are front-line in evaluating patients with stroke in the community. Their ability to correctly identify stroke influences downstream management decisions. We sought to use a large national database of prehospital clinical data to determine risk factors associated with missed EMS stroke identification.

**METHODS:** Retrospective study examining EMS evaluation of adults with Emergency Department (ED) stroke diagnosis. We leveraged the ESO Data Collaborative research dataset containing EHR data from 2019-2022 that has a subset of encounters with linked hospital diagnostic codes. Our primary outcome was the presence of an EMS diagnosis of stroke. We evaluated the association between demographic and clinical variables with EMS stroke identification using Pearson χ2 test for demographic variables and multivariable GLM for clinical variables with adjustment for demographic variables.

**RESULTS:** We identified 34,504 EMS encounters for patients with ED stroke diagnosis. Of these, 11,077 (32.1%) strokes had missed EMS stroke identification and instead had an EMS impression of “Generalized Weakness” (25.9%), “Altered Level of Consciousness” (24.9%), and “Dizziness” (7.2%). Patients more likely to have missed prehospital stroke identification were of Black race (p=0.0001) and Hispanic ethnicity (p=0.0001). Clinical variables associated with higher risk of missed EMS stroke identification were suspected alcohol or drug use (RR 1.48, 95% CI 1.37-1.59), low GCS (RR 1.17, 95% CI 1.10-1.24), tachycardia (RR 1.05, 95% CI 1.01-1.09), and hypotension (RR 1.47, 95% CI 1.34-1.61).

**CONCLUSIONS:** Approximately 1-in-3 patients transported by EMS did not have their stroke identified in the prehospital setting. Factors associated with lower odds of missed EMS stroke identification provide a starting point for future performance improvement initiatives.

## Background

Emergency medical services (EMS) provide care for over 250,000 patients hospitalized with stroke in the US annually. Prehospital stroke recognition and pre-arrival alerting are linked with improved rates of time-sensitive treatment with thrombolysis and thrombectomy.^1–4^ Conversely, lack of EMS stroke recognition may result in transport to a facility not equipped to treat patients with stroke and may result in delays in care.

Prior single-center studies estimate that 30% of strokes are not identified during EMS evaluation.^3,5^ However, these studies were conducted before thrombectomy was established as the standard of care for patients with stroke due to a large vessel occlusion. Work has been done to improve EMS training on stroke recognition and the use of prehospital stroke screening scales,^6^ but whether these efforts have improved rates of EMS stroke identification is unclear.^7,8^

Moreover, the clinical and social factors associated with an increased risk of missed stroke diagnosis during EMS evaluation are unknown, despite prior studies showing associated delayed treatment and worse outcomes in people from historically underserved or marginalized backgrounds.^9–11^ Prior studies are largely limited to patients presenting to urban academic hospitals with findings that may not be generalizable to non-academic or rural hospitals. This study aimed to quantify the proportion of EMS transported patients who were diagnosed with strokes that were not identified during prehospital evaluation. Secondarily we aimed to identify the clinical and sociodemographic factors associated with missed prehospital stroke identification using a geographically broad national dataset of linked EMS and hospital records.

## Methods

### Study Design and Setting

We conducted a retrospective analysis of adults who were transported by EMS after a 911 activation and were subsequently diagnosed with stroke during the emergency department (ED) evaluation. We used the ESO Data Collaborative public-use research dataset. ESO is a leading provider of pre-hospital Electronic Health Record (EHR) systems for EMS clinical documentation in the US.^12^ ESO EHR collects data in accordance with the National EMS Information System. Data elements in the prehospital EHR include information related to EMS dispatch and the prehospital clinical encounter. EMS clinicians document encounters using prespecified data fields, which include diagnostic impressions and whether a ‘stroke treatment protocol’ was used. The ESO Data Collaborative consists of all records from agencies who have agreed to share their de-identified EHR data for the purposes of research. Annually, a de-identified dataset is constructed with all records from participating agencies. This dataset includes EMS responses in every region of the country. A subset of destination facilities (i.e. hospitals) participate in the ESO Health Data Exchange (HDE) which allows prehospital data to be directly linked with hospital EHR data using HL7 messaging including ICD-10 diagnosis codes and discharge dispositions. This study was approved by the institutional review board of the University of California, San Francisco.

### Study Population

We identified adult patients aged 18 years or older who were transported to a hospital following a 9-1-1 EMS response, and were diagnosed with an acute ischemic stroke in the ED between January 1, 2019 and December 31, 2022. A diagnosis of acute ischemic stroke in the ED was defined as having an ICD-10 primary discharge diagnosis code of cerebral infarction (I63.x).

### Outcome

The primary outcome was missed EMS stroke identification. Missed EMS stroke identification was defined as no recorded diagnostic impression of stroke during EMS evaluation and no indication of stroke protocol use.

This definition assumes that the EMS diagnostic impression is an accurate reflection of whether or not an EMS clinician suspects a patient is experiencing a stroke. To determine whether this was a reasonable assumption, we examined the narrative history recorded by the EMS clinician from a random sample of 300 encounters in the cohort and found that a minority of patients were misclassified using this definition (Appendix; Supplemental Tables 1 & 2).

### Measurements

Demographics, diagnostic impressions, clinical information (e.g. vital signs and Glasgow Coma Scale [GCS]), and the EMS agency treatment protocol associated with each encounter are entered into the prehospital EHR by EMS clinicians as part of the required documentation following a 911 call. We evaluated patient age, sex, race, ethnicity, Census region, urbanicity of the community where the encounter took place, the first recorded prehospital vital sign measurements, GCS, and whether the EMS clinician suspected alcohol or substance use. Age was divided into ordinal groups of <40, 40–59, 60–79, and >80 years. Race was recorded by the EMS clinicians and categorized as White, Black or African American, Asian or Pacific islander, or Other/unknown. Ethnicity was categorized as Hispanic or non-Hispanic; these were collected because of previously-reported disparities in stroke care for Hispanic patients.^9,11^ Social vulnerability index (SVI) – a measure of socioeconomic factors associated with adverse community-level hazards and stressors – was categorized into four quartiles from least to most vulnerable. Urbanicity was determined by Rural-Urban Commuting Area Codes and categorized as: Metropolitan (population > 49,999) or Non-metropolitan (population < 50,000). Census regions were categorized as Northeast, South, Midwest; or West. GCS and vital sign measurements were divided into ordinal groups in alignment with conventionally accepted normal and abnormal ranges for adults to ease the interpretation of effect estimates.

### Statistical Analysis

We calculated the proportion of patients with ED-diagnosed stroke where there was missed EMS identification of stroke during prehospital evaluation. We identified sociodemographic and clinical characteristics associated with missed EMS stroke identification using Pearson χ2 test for the unadjusted analyses and binomial family and log link generalized linear models for the adjusted analyses. The models allowed us to examine the association of initial vital sign measurements, level of consciousness, and suspected alcohol or drug use, with the risk of missed EMS stroke identification. We calculated unadjusted and adjusted estimates separately for each exposure, adding patient age, sex, race, GCS, and urbanicity in the adjusted analyses.

Because ED encounters in rural areas are not well represented in existing literature, we sought to determine whether urbanicity modifies the likelihood and risk factors of having missed EMS stroke identification. To do this, we repeated the models after stratifying by whether the encounter originated in an urban or rural environment. All reported risk ratios (RRs) are from adjusted models unless otherwise specified. Statistical analyses were performed using Stata (version 15.1, StataCorp, College Station, TX).

## Results

We analyzed 34,504 EMS encounters for patients that were diagnosed with stroke in the ED. Most (94.7%) encounters that occurred in a metropolitan area. Approximately half were female (51.3%). Two thirds (64.3%) of patients were White, and 16.9% were Black and 8.1% had documentation of Hispanic ethnicity.

There were 11,077 (32.1%) encounters which did not have a prehospital diagnostic impression of stroke by EMS evaluation. The most common EMS diagnostic impressions for those without stroke recognition were “Generalized Weakness” (25.9%), “Altered Level of Consciousness” (24.9%), “Dizziness” (7.2%), “Other Cardiovascular” (5.7%), and “Pain” (3.7%) (Table 1).

**Table 1:**
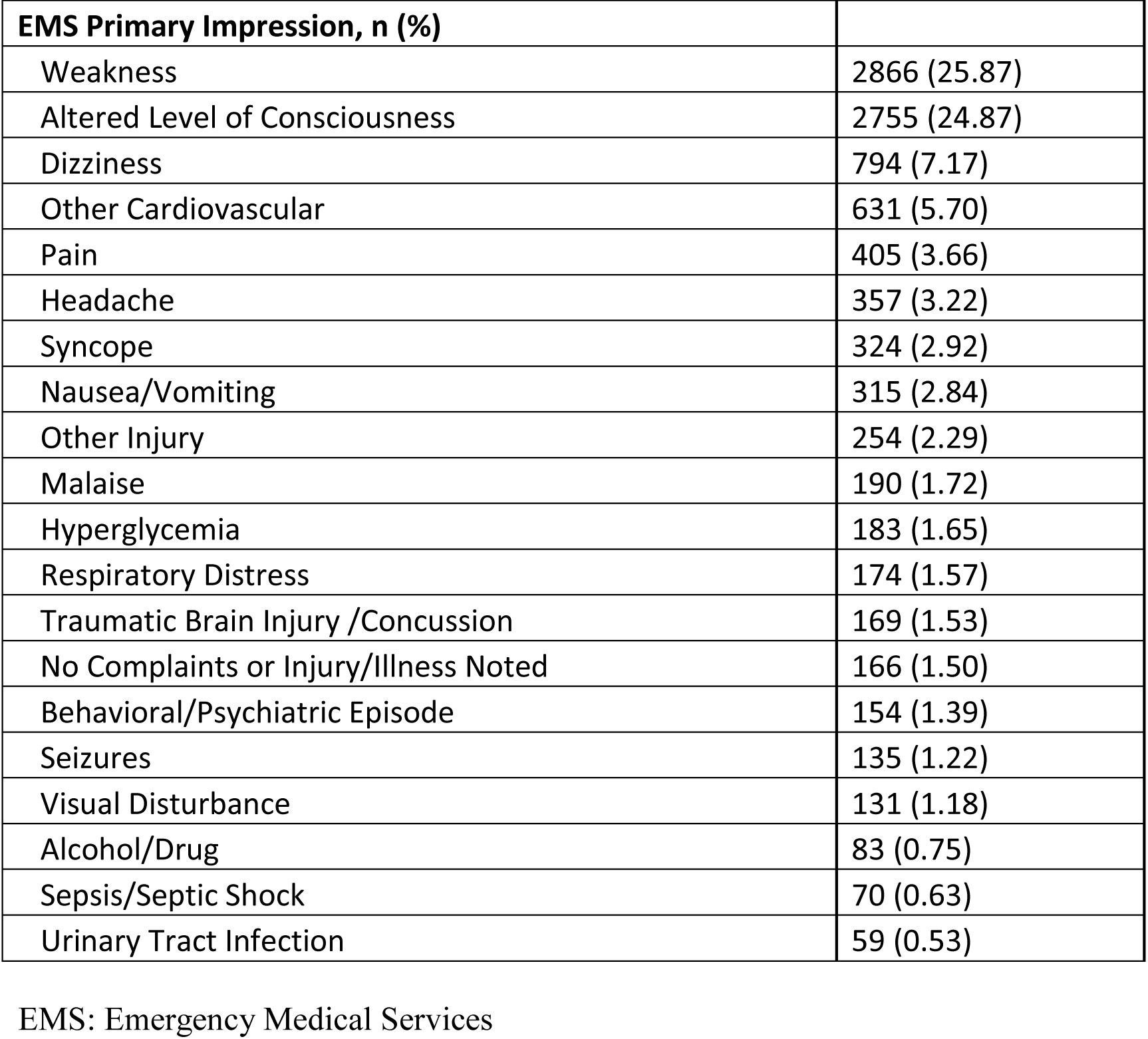
EMS primary impressions of patients without stroke recognition.

| EMS Primary Impression, n (%) |  |
| --- | --- |
| Weakness | 2866 (25.87) |
| Altered Level of Consciousness | 2755 (24.87) |
| Dizziness | 794 (7.17) |
| Other Cardiovascular | 631 (5.70) |
| Pain | 405 (3.66) |
| Headache | 357 (3.22) |
| Syncope | 324 (2.92) |
| Nausea/Vomiting | 315 (2.84) |
| Other Injury | 254 (2.29) |
| Malaise | 190 (1.72) |
| Hyperglycemia | 183 (1.65) |
| Respiratory Distress | 174 (1.57) |
| Traumatic Brain Injury /Concussion | 169 (1.53) |
| No Complaints or Injury/Illness Noted | 166 (1.50) |
| Behavioral/Psychiatric Episode | 154 (1.39) |
| Seizures | 135 (1.22) |
| Visual Disturbance | 131 (1.18) |
| Alcohol/Drug | 83 (0.75) |
| Sepsis/Septic Shock | 70 (0.63) |
| Urinary Tract Infection | 59 (0.53) |

EMS: Emergency Medical Services

A larger proportion of Black patients had missed EMS stroke identification compared to White patients (34.9% vs 31.4%, p<0.001). Similarly, a larger proportion of Hispanic patients had missed EMS stroke identification compared to non-Hispanic patients (36.5% vs 31.7%, p<0.001). A larger proportion of patients with high SVI had missed EMS stroke identification compared to patients with low SVI (28.2% vs 21.4%, p<0.001) Stroke patients with missed EMS stroke identification during EMS evaluation were otherwise similar with respect to age, sex, and urbanicity. (Table 2).

**Table 2.** Patient Demographic Characteristics.

|  | Overall | EMS Impression |  |  |
| --- | --- | --- | --- | --- |
|  |  | Stroke | Not Stroke | p-value* |
| N | 34504 | 23427 | 11077 |  |
| Age, mean (SD) | 71.6 (14.3) | 71.9 (14.2) | 70.9 (14.5) | <0.0001 |
| Female, n (%) | 17652 (51.3%) | 11938 (51.1%) | 5714 (51.8%) | 0.29 |
| Race, n (%) |  |  |  | <0.0001 |
| Asian, Native Hawaiian,<br>or Pacific Isl. | 645 (1.9%) | 429 (1.8%) | 216 (1.9%) |  |
| Black or African<br>American | 5823 (16.9%) | 3789 (16.2%) | 2034 (18.4%) |  |
| White | 22185 (64.3%) | 15211 (64.9%) | 6974 (63.0%) |  |
| Other, unknown, or<br>American Native | 5851 (17.0%) | 3998 (17.1%) | 1853 (16.7%) |  |
| Hispanic ethnicity, n (%) | 2351 (8.1%) | 1493 (7.6%) | 858 (9.0%) | <0.0001 |
| Social Vulnerability Index<br>Quartile, N (%) |  |  |  | <0.0001 |
| Least Vulnerable | 8307 (24.1%) | 5944 (25.4%) | 2363 (21.4%) |  |
| Quartile 2 | 9083 (26.4%) | 6319 (27.0%) | 2767 (25.0%) |  |
| Quartile 3 | 8625 (25.0%) | 5810 (24.8%) | 2815 (25.4%) |  |
| Most Vulnerable | 8452 (24.5%) | 5333 (22.8%) | 3119 (28.2%) |  |
| Urbanicity, n (%) |  |  |  | 0.22 |
| Non-Metro Area | 1834 (5.3%) | 1221 (5.2%) | 613 (5.5%) |  |
| Metro Area | 32617 (94.7%) | 22168 (94.8%) | 10449 (94.5%) |  |
| Census Region, n (%) |  |  |  | <0.0001 |
| Midwest | 2052 (18.7%) | 1380 (18.4%) | 672 (19.4%) |  |
| Northeast | 716 (6.5%) | 559 (7.4%) | 157 (4.5%) |  |
| South | 5376 (49.0%) | 3631 (48.3%) | 1745 (50.4%) |  |
| West | 2827 (25.8%) | 1941 (25.8%) | 886 (25.6%) |  |

\*p-values calculated using chi-square for categorical variables and t-test for means

### Clinical Risk Factors for Missed Prehospital Stroke Identification

Suspected alcohol or drug use, GCS score, and vital sign abnormalities were associated with risk of missed EMS stroke identification. Suspected alcohol or drug use was associated with a 48% increased risk of missed EMS stroke identification (RR 1.48, 95% CI, [1.37 to 1.59]). Severe depression in consciousness (GCS 3-8) was associated with a 17% increased risk of missed EMS stroke identification (RR 1.17, [1.10 - 1.24]). Tachycardia (RR 1.05, [1.01 – 1.09]), hypotension (RR 1.22, [1.15 – 1.30]), and bradypnea (RR 1.44, [1.19 – 1.74]) were also associated with an increased risk of missed EMS stroke identification (Table 3).

**Table 3.** Patient Clinical Characteristics.

|  | EMS Impression |  | Risk of missed EMS stroke identification |  |
| --- | --- | --- | --- | --- |
|  | Stroke<br>(n = 23427) | Not Stroke<br>(n = 11077) | RR (95% CI)* | aRR (95% CI)** |
| Alcohol and Drug Use |  |  |  |  |
| None indicated | 23013 (98.2%) | 10696 (96.6%) | [ref] | [ref] |
| Indicated | 414 (1.8%) | 381 (3.4%) | 1.51 (1.40, 1.63) | 1.48 (1.37, 1.59) |
| GCS |  |  |  |  |
| 8 or less | 1021 (4.5%) | 658 (6.1%) | 1.17 (1.10, 1.24) | 1.17 (1.10, 1.24) |
| 9 - 12 | 3696 (16.4%) | 1051 (9.8%) | 0.66 (0.63, 0.70) | 0.67 (0.63, 0.70) |
| 13 - 15 | 17905 (79.1%) | 9018 (84.1%) | [ref] | [ref] |
| Heart Rate |  |  |  |  |
| <60 (bradycardia) | 1293 (5.5%) | 590 (5.4%) | 0.99 (0.92, 1.06) | 1.01 (0.94, 1.08) |
| 60-100 (normal) | 17622 (75.5%) | 8216 (74.5%) | [ref] | [ref] |
| > 100 (tachycardia) | 4418 (18.9%) | 2215 (20.1%) | 1.05 (1.01, 1.09) | 1.05 (1.01, 1.09) |
| Systolic Blood Pressure |  |  |  |  |
| <90 (hypotension) | 228 (1.0%) | 241 (2.2%) | 1.48 (1.35, 1.63) | 1.47 (1.34, 1.61) |
| 90-140 (normal) | 6841 (29.4%) | 3621 (32.9%) | [ref] | [ref] |
| >140 (hypertension) | 16229 (69.7%) | 72151 (64.9%) | 0.88 (0.86, 0.91) | 0.87 (0.84, 0.90) |
| Diastolic Blood Pressure |  |  |  |  |
| <60 (hypotension) | 1120 (4.9%) | 750 (6.9%) | 1.21 (1.14, 1.28) | 1.22 (1.15, 1.30) |
| 60-90 (normal) | 12495 (54.3%) | 6189 (56.9%) | [ref] | [ref] |
| >90 (hypertension) | 9408 (40.9%) | 3931 (36.2%) | 0.89 (0.86, 0.92) | 0.87 (0.84, 0.90) |
| Respiratory Rate |  |  |  |  |
| <8 | 58 (0.3%) | 56 (0.5%) | 1.54 (1.27, 1.85) | 1.44 (1.19, 1.74) |
| 8-20 | 20223 (88.8%) | 9502 (88.1%) | [ref] | [ref] |
| >20 | 2492 (10.9%) | 1233 (11.4%) | 1.04 (0.99, 1.09) | 1.05 (1.00, 1.10) |
| Oxygen Saturation |  |  |  |  |
| < 89 | 933 (4.1%) | 476 (4.5%) | 1.05 (0.98, 1.13) | 1.09 (1.01, 1.18) |
| 89 + | 21561 (95.9%) | 10215 (95.5%) | [ref] | [ref] |
| Fever |  |  |  |  |
| <95 | 51 (0.6%) | 38 (0.7%) | 1.17 (0.92, 1.49) | 1.24 (0.98, 1.56) |
| 95-100.4 | 9043 (98.3%) | 5197 (96.7%) | [ref] | [ref] |
| >100.4 | 106 (1.2%) | 138 (2.6%) | 1.55 (1.39, 1.73) | 1.54 (1.37, 1.72) |

\*Unadjusted generalized linear models (GLMs) were run for each clinical characteristic as the single clinical exposure
\*\*Adjusted GLMs were calculated to include the single exposure along with covariates of patient age, sex, race, GCS, and urbanicity.
ED: Emergency Department, EMS: Emergency Medical Services, GCS: Glasgow Coma Score,
GLM: Generalized Linear Model, RR: Relative Risk, aRR: adjusted Relative Risk.

After stratifying the main analysis by whether the encounter occurred in a metropolitan area, suspected alcohol or drug use, GCS, and vital sign abnormalities remained associated with an increased risk of missed EMS stroke identification in both settings (Supplemental Table 3).

## Discussion

This is the largest cohort study of the EMS stroke diagnostic sensitivity in the United States; we found that nearly one-third of strokes were missed during EMS evaluation. Our findings are consistent with estimates obtained before mechanical thrombectomy was established as standard therapy for large vessel occlusive stroke.^13,14^ This suggests that the rates of missed EMS stroke identification remain largely unchanged despite efforts to improve stroke recognition in the prehospital setting.

Black and Hispanic patients had a significantly higher risk of missed EMS stroke identification. Prehospital notification is already shown to be less likely to be used for Black and Hispanic patients with stroke.^3,5^ Our findings suggest that the failure to recognize stroke in the field may contribute to reduced rates of hospital pre-arrival stroke notification in Black and Hispanic patients. We additionally found that a report of alcohol or substance use increased the risk of missed EMS stroke identification. This novel finding suggests that alcohol use may distract clinicians from developing a suspicion of stroke.

This is the first study to examine prehospital clinical characteristics that modulate the risk of missed EMS stroke identification: we found that prehospital hypotension, tachycardia, and loss of consciousness were associated with an increased risk of missed EMS stroke identification. This may be related to incorrect clinical assumptions that hypotension and tachycardia are reflective of a separate pathophysiological process – such as sepsis – that misleads the clinician away from a stroke diagnosis.^15,16^ Patients with a low GCS (3-8) also was associated with a higher risk of missed EMS stroke identification compared to patients with near normal GCS. The inability to obtain a nuanced neurological assessment in nearly comatose patients likely obscures ascertainment of stroke. While it is unlikely these patients are managed with any less urgency than patients who have stroke recognized by EMS, it is possible that patient triage and hospital destination decisions differ between the two groups. Further research is needed to compare management between patients where the stroke was recognized versus missed in those who present with disorders of consciousness.

We found that *half* of patients who had missed EMS stroke identification were given a diagnostic impression of generalized weakness or altered level of consciousness. Attempts to improve rates of EMS stroke identification and hospital prenotification have focused on educational interventions targeted to EMS clinicians evaluating patients already suspected to have a stroke.^7,8^ While shown to be useful in rates of EMS stroke identification, the benefit of one brief educational module was not sustained after 3 months;^17^ a separate enhanced paramedic stroke assessment method in patients with suspected stroke actually lengthened the time of prehospital care episodes and delayed thrombolysis.^18^ Our finding suggests that educational efforts should focus on expanding the use of prehospital neurological assessments to all patients who present with weakness and altered level of consciousness.

Limitations to this study include the reliance on an EMS diagnostic impressions of stroke and protocols used as a proxy for when EMS clinicians suspected stroke. This study did not examine how EMS stroke identification influences downstream management – such as hospital pre-arrival notification by EMS, hospital destination decisions, and in-hospital stroke care. Finally, this study demonstrates but does not explain racial and ethnic disparities in EMS stroke identification, and whether this is reflective of individual bias occurring on the part of the EMS clinician or structural factors that drive disparate health characteristics in racialized populations at-large.

Our findings highlight that a large proportion of patients with stroke do not have the stroke identified by EMS. Patients who identify as Black and Hispanic are disproportionately affected, and poor mental status and vital sign abnormalities not traditionally associated with stroke are also associated with an increased risk of missed EMS stroke identification. Educational interventions to improve EMS stroke identification can focus on maintaining a suspicion of stroke in diverse clinical contexts to improve stroke identification.

## Data Availability

Data are available upon reasonable request to ESO Inc.

https://www.eso.com/

## Disclosures

Dr. Caffarelli is named as sole inventor on a patent on the use of electroencephalography for stroke diagnosis with no associated licensing agreements.

Dr. Kamel has a PI role in the ARCADIA trial, which received in-kind study drug from the BMS-Pfizer Alliance for Eliquis and ancillary study support from Roche Diagnostics; a Deputy Editor role for *JAMA Neurology*; clinical trial steering/executive committee roles for the STROKE-AF (Medtronic), LIBREXIA-AF (Janssen), and LAAOS-4 (Boston Scientific) trials; consulting or endpoint adjudication committee roles for AbbVie, AstraZeneca, Arthrosi Therapeutics, Boehringer Ingelheim, Eli Lilly, and Novo Nordisk; and household ownership interests in TETMedical, Spectrum Plastics Group, and Ascential Technologies.

Dr. Kim is an Associate Editor of NEJM and Journal Watch: Neurology

Dr. Guterman receives personal compensation from JAMA Neurology and stock from REMO Health, which are unrelated to the submitted work.

## ABBREVIATIONS

CI: Confidence Interval
EHR: Electronic Health Record
EMS: Emergency Medical Services
ED: Emergency Department
GCS: Glasgow Coma Scale
HDE: Health Data Exchange
RR: Relative Risk
SVI: Social Vulnerability Index

